# The Solute Carrier Family 26 Member 9 Is a Modifier of the Rapidly Progressing Cystic Fibrosis Associated with F508del CFTR Mutations

**DOI:** 10.1101/2024.01.04.23300546

**Authors:** Shiyu Luo, Stuart Rollins, Klaus Schmitz-Abe, Amy Tam, Qifei Li, Jiahai Shi, Jasmine Lin, Ruobing Wang, Pankaj B. Agrawal

**Affiliations:** Division of Neonatology, Department of Pediatrics, University of Miami Miller School of Medicine and Holtz Children’s Hospital, Jackson Health System, Miami, FL 33136, USA; Division of Genetics and Genomics; The Manton Center for Orphan Disease Research, Boston Children’s Hospital and Harvard Medical School, Boston, MA 02115, USA; Division of Pulmonary Medicine, Boston Children’s Hospital; Department of Medicine, Harvard Medical School; Department of Biochemistry, Yong Loo Lin School of Medicine, National University of Singapore, Singapore, Singapore; Center for Regenerative Medicine of Boston University and Boston Medical Center; Boston, MA 02115, USA

**Keywords:** SLC26A9, cystic fibrosis, modifier gene, deltaF508

## Abstract

Cystic fibrosis (CF) is an autosomal recessive disease caused by mutations to the CF transmembrane conductance regulator (*CFTR*). Symptoms and severity of the disease vary shown that modifier genes influence disease severity and clinical course. We previously reported epithelial sodium channel (ENaC) genes as modifiers of disease severity in long-term non-progressors sharing deltaF508 homozygous for *CFTR* genotype. Here we describe the opposite, modifier genes that may be associated with rapidly progressing CF (RPCF) in a cohort of patients with the shared deltaF508 homozygous genotype. We have identified three rare missense *SLC26A9* variants in four individuals (out of six) deemed to have RPCF: c.229G>A; p.G77S (present in two patients), c.1885C>T; p.P629S and c.2546G>A; p.R849Q. By analyzing publicly available single cell sequencing dataset from LungMAP, we revealed that both *SLC26A9* and *CFTR* mRNA are highly enriched in Alveolar type 2 (AT2) cells, with a few (greater than 1%) in respiratory airway secretory (RAS) cells and ionocytes. Structural modeling suggests deleterious effects of these mutations as they are in critical protein domains which might affect the ion transportation capability of SLC26A9. The enrichment of rare and potentially deleterious *SLC26A9* mutations in patients with RPCF suggests *SLC26A9* is a modifier gene associated with RPCF.

## Introduction

Cystic fibrosis (CF) is a chronic and multisystem disease affecting more than 30 000 individuals in the US and approximately 89 000 worldwide (1). CF is caused by recessive mutations in the CF transmembrane conductance regulator gene (*CFTR*), and of people with CF (pwCF) in the US, approximately 85.5% carry the variant F508del (1). CFTR is located in the apical membrane of epithelia and regulates the sodium and chlorine ion transport and subsequent electrolyte and water balances in multiple organs (2). Loss of CFTR is associated with a spectrum of disease related to exocrine dysfunction, including chronic respiratory bacterial infections associated with obstructive airway disease, pancreatic insufficiency with malnutrition, cystic fibrosis-related diabetes (CFRD), cystic fibrosis-related liver diseases (CFRLD), and reduced life expectancy. Its respiratory phenotype is highly penetrant, with a typical loss of lung function of ∼2.5%/year in the forced expiratory volume in 1 second (FEV_1_), translating to a median life expectancy of 47 to 53 years despite new effective therapies and advanced multidisciplinary care (1, 3, 4).

The disease severity in CF varies greatly: pwCF with similar *CFTR* variants and colonized with similar microbiota, may have highly variable clinical manifestations (5). It is recognized that genetic variants located outside the CFTR locus, "modifier genes", influence the clinical expression of the disease and therapeutic efficacy (6, 7). Although many genes have been proposed as potential modifiers of CF severity, only a handful have withstood the test of replication or have strong functional evidence. These include the mucin MUC4/MUC20 locus, Ets Homologous Factor (EHF)/Apaf-1 interacting protein (APIP) locus, genes encoding epithelial sodium channel (ENaC), solute carrier family 9 member A3 (SLC9A3), solute carrier family 6 member 14 (SLC6A14), Alpha1-antitrypsin (SERPINA1), and transcription factor 7-like 2 (TCF7L2) with varying level of modifier effects (7-14). Exome or genome sequencing (ES/GS) has become a powerful and effective strategy for the discovery of genes underlying Mendelian disorders. However, use of ES/GS to identify variants associated with complex traits has been more challenging, partly because of the need for large sample size to adequately power the study. One strategy to improve identification of modifiers is to sequence individuals who are at extreme ends of a phenotype distribution. Because the frequency of alleles that contribute to the trait are enriched in one or both phenotype extremes, a modest sample size can be used to identify novel candidate genes and/or alleles (10, 15, 16).

By utilizing this strategy, we have previously identified the epithelial sodium channel as a modifier of the long-term nonprogressive phenotype (LTNP) associated with homozygous F508del CFTR mutations (10). Here, we use an unbiased genome-wide genetic burden test to analyze ES data from patients affected by rapidly progressing CF (RPCF) to identify candidate modifier genes. We have identified SLC26A9 as one of the key CF modifier genes and report three missense variants in four patients (out of six) with RPCF phenotype. We explore the publicly available single cell sequencing dataset from LungMAP and visualize the expression of *SLC26A9* in comparison with *CFTR*. In addition, structural modeling was performed to evaluate the deleterious effects of these mutations.

## Materials & Methods

### Participants

From a large group of CF patients carrying homozygous F508del mutations, we have previously identified eight LTNP individuals who had a long survival time and stable lung-function trajectory (10). In this study, we have identified six rapidly progressing CF (RPCF) patients who had fast-deteriorating lung function, needing lung transplant or dying early before the age of 18. Sequencing data from the control group (n = 3076) was obtained from a cohort of patients not affected by CF but with different clinical phenotypes, such as autism, microcephaly, etc. as well as their parents. Both case and control data were obtained from patients at our institution with consent according to approved IRB protocols. This study has been approved by the Institutional Review Board (IRB) of Boston Children’s Hospital affiliated with Harvard Medical School (IRB protocol no.:10-02-0253). All patient/participant consent has been obtained and the appropriate institutional forms archived.

### Exome Sequencing and Data Analysis

Blood samples were collected and processed for DNA extraction. DNA was sent for exome sequencing to GeneDx (Gaithersburg, MD). WES and data analysis were performed as previously described using VExP (17). Data from FASTQs or BAMs/CRAMs was aligned or realigned to GRCh37 using BWA MEM and further processed using SAMtools and Picard. Variants were called using GATK. Variant calls were inputted into an in-house program that flags and scores mutations from each patient sample using a neural network and takes as input: 1) read quality; 2) population statistics from various databases, such as gnomAD, and 1000 Genome Project; 3) constraint scores from gnomAD; 4) results from various predictors of mutation effect, such as PolyPhen, SIFT, and CADD; and 5) results from various predictors of spliceogenicity, such as SpliceAI and SPiCE.

### Gene-based Burden Test and Analysis

Intuitively, we filtered for variants to enrich for genes most likely to act as modifier genes and compared how often the gene was mutated in patients affected by RPCF compared to those who were not. Taking results from VExP, candidate mutations (MAF < 1.5%) were segregated into Dominant and Recessive models and collapsed gene-wise in three bins: 1) missense variants 2) loss-of-function (as defined in gnomAD) variants 3) combination of loss-of-function and missense variants.

We then performed a Fisher’s exact test for each of these three bins for each gene between samples from patients affected by RPCF or LTNP against patients not affected by CF. Taking these results, we focused on candidate modifier genes that: a) followed a Dominant model; b) had at least 80% of variant calls passing quality checks; c) were statistically significant for RPCF (uncorrected Fisher p-value < 0.001); and d) were NOT significant for LTNP (uncorrected Fisher p-value > 0.05).

We further reviewed these candidate modifier genes for RPCF using phenotypic data, expression, gene function, and constraint data found on publicly available databases such as OMIM, GTEx, UniProt, the UCSC Genome Browser, and gnomAD, as well as publications found on PubMed.

### Molecular Modeling

The SLC26A9 protein structure model is built on its structure (PDB code: 7CH1) determined by Cryo-Electron Microscopy in the form of a homodimer (18). The protein structure illustration and amino acid mutations are generated by PyMOL (The PyMOL Molecular Graphics System, version 2.0 Schrödinger).

## Results

### Patient Profiles

We identified six (6) pediatric patients from our institution who were affected by cystic fibrosis carrying two copies of the *CFTR* p.F508del mutation and met our definition of RPCF. Of these 6 patients, four were female and two were male. All were pancreatic insufficient and have had *Pseudomonas aeruginosa*, among other microbiology, identified in their lung cultures. A majority also had records of high sweat chloride, low BMI, and CF-related sequelae, such as CF-related diabetes and liver disease among other comorbidities (Table 1). Three patients have received lung transplants and one of them died a couple of months after transplant due to complications. Two other patients have died between age 10 and 20 years old. One patient who didn’t receive lung transplant currently receives elexacaftor/tezacaftor/ivacaftor (ETI) therapy. **Table 1** lays out demographic and clinical characteristics for these patients. **Figure 1A** shows representative computed tomography (CT) scan images from two RPCF patients, and **Figure 1B** presents the evaluation of all six patients’ lung function, based on their forced expiratory volume in 1 second (FEV1) value which represents the maximum amount of air that a person can forcibly expel during the first second following maximal inhalation.

**Figure 1:**
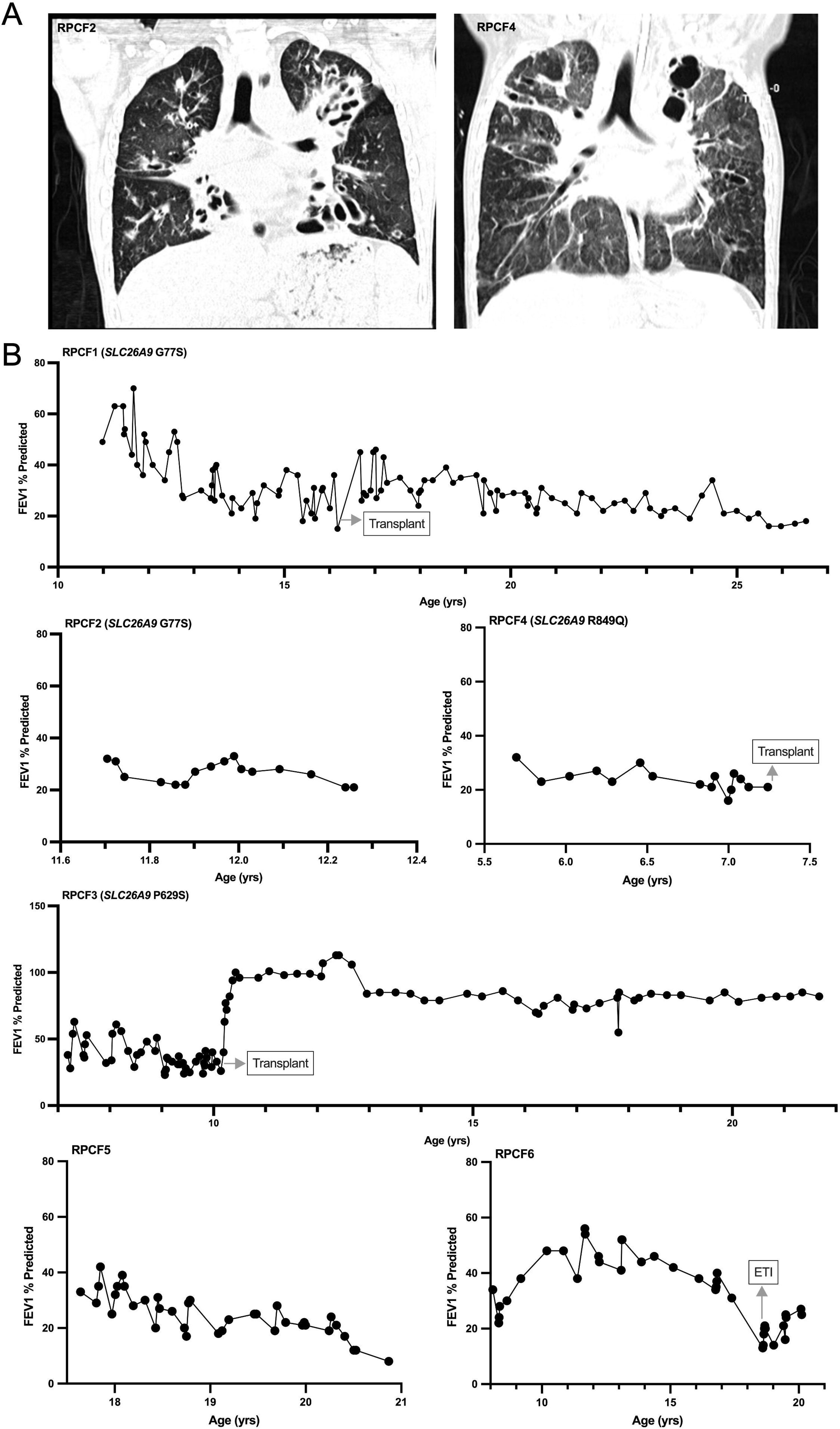
Representative CT scan images **(A)** and longitudinal analysis of FEV_1_ % predicted **(B)** for F508del RPCF patients. **(A)** The left panel shows a CT scan from RPCF2, showing diffuse bilateral bronchiectasis and bronchiolar wall thickening. Right panel is from RPCF4, when she was intubated with extensive, severe bronchiectasis with endobronchial secretions, air-trapping and scarring, extensive mediastinal lymphadenopathy and extensive tree-in-bud opacities throughout the lungs. Please note that the labeling Pt no. were not known to anyone outside the research group.

**Table 1:** Clinical Phenotypes of the Six Rapidly Progressing Cystic Fibrosis (RPCF) patients.

| Pt. no | Gender | Age range (years) | SLC26A9 Mutation | Intervention: age range (years) | Microbiology | Sweat Chloride | BMI | Pancreatic Function | Other Comorbidities |
| --- | --- | --- | --- | --- | --- | --- | --- | --- | --- |
| RPCF1 | M | 26-30 | SLC26A9 G77S | Transplant: 16-20 | PsA<br>MSSA<br>Steno<br>Burkholderia gladioli | n/a | LBI: 15.6 (20.4 %ile)<br>RBI: 20.1 (41 %ile) | PI | Severe ABPA<br>Asthma<br>CFRD<br>GERD<br>Gastrointestinal (GI) bleeding<br>Obstructive sleep apnea<br>Developmental delay<br>Myotonic dystrophy |
| RPCF2 | F | Death: 11-15 | SLC26A9 G77S & P322P | No | PsA<br>MSSA<br>B. cepacia | n/a | LBD: 13 (0.17 %ile)<br>RBD: 14 (3.4 %ile) | PI | n/a |
| RPCF3 | M | 21-25 | SLC26A9 P629S | Transplant: 6-10 | PsA<br>MSSA<br>Steno | n/a | LBI: 16 (40.7 %ile)<br>RBI: 16.4 (44 %ile) | PI | Asthma<br>ABPA<br>CFRD |
| RPCF4 | F | Death: 6-10 | SLC26A9 R849Q | Transplant: 6-10 | PsA<br>Aspergillus<br>Mycobacterium abscessus | 104, 102 | LBD: 14.4 (26 %ile)<br>RBD: 17.2 (78.7 %ile) | PI | n/a<br>had burkholderia |
| RPCF5 | F | Death: 16-20 |  | No but advised transplant before age 18 | PsA<br>MAC<br>Aspergillus | n/a | LBD: 17.7 (24 %ile)<br>RBD: 19.8 (25 %ile) | PI | CFRD<br>CFRLD<br>Chronic sinus disease<br>Esophageal varices<br>GI bleed<br>Depression<br>Anxiety |
| RPCF6 | F | 16-20 |  | ETI: 16-20 | PsA<br>MSSA | 84,92 - >ETI-> 30,26 | LBI: 13.1 (0.03 %ile)<br>RBI: 15.4 (0.03 %ile) | PI | Ischemic stroke |

Definition of abbreviations: ABPA, allergic bronchopulmonary aspergillosis; CFRD, cystic fibrosis–related diabetes; CFRLD, cystic fibrosis–related liver diseases; F, female; GERD, gastroesophageal reflux disease; M, male; MAC, Mycobacterium avium complex; MSSA, methicillin-sensitive Staphylococcus aureus; PI, pancreatic insufficiency; PsA, Pseudomonas aeruginosa; Steno, Stenotrophomonas maltophilia. LBD, lowest before death; LBI, lowest before intervention; RBD, right before death; RBD, right before death.

### Gene-based Burden Analysis

These six patients with RPCF provided samples for ES, and we further investigated by performing an unbiased burden test for all coding genes identified in the human genome using patient and control sequencing data at our institution. We focused on the dominant models and performed Fisher’s exact test between our cohort of six RPCF individuals against 3076 individuals with no CF who served as controls, as well as a parallel analysis between the eight individuals affected by LTNP and same controls. From these results, we selected for non-synonymous variants in genes with p-value < 0.001 for RPCF, but not significant (p-value > 0.05) for LTNP. This process yielded 53 candidate genes (**supplementary Table S1**) that are most likely to be functional (non-synonymous variants) and associated with RPCF (p-value significant for RPCF and not significant for LTNP).

Then we reviewed these genes using phenotypic, expression, gene function, and constraint data found on publicly available databases, as well as publications found on PubMed. *SLC26A9* most significantly associated with RPCF with an uncorrected p-value of 1.10E-05 for RPCF (*SLC26A9* missense variants were carried by 4 out of 6 RPCF patients, 0 out of 8 LTNP patients and none of 3076 controls).

Although we focused on dominant models in this study to identify candidate modifier genes, we also performed a parallel burden analysis on recessive models to compare the patients affected by CF against patients not affected by CF. As expected, the *CFTR* gene appeared as the most statistically associated gene. *CFTR* LoF + missense variants had an uncorrected p-value of 1.6E-15 for our patients affected by RPCF.

### LungMAP Expression Analysis of SLC26A9 in Comparison with CFTR Using Human and Mouse Single Cell RNA Sequencing Datasets

Genotype-Tissue Expression (GTEx) analysis showed that a variety of tissues, such as digestive system, lungs, and skin, express both *CFTR* (ENSG00000001626.14) and *SLC26A9* (ENSG00000174502.18) at various levels as measured through bulk RNA sequencing of non-diseased tissue from nearly 1000 participants (19). As this study focus on a potential genetic modifier role of SLC26A9 on the expression of cystic fibrosis lung phenotype, we explored its expression along with CFTR in different cell types using LungMAP (20, 21). In the human lung tissues(22), *SLC26A9* mRNA is mostly enriched in Alveolar type 2 (AT2), Pulmonary neuroendocrine (PNEC), and respiratory airway secretory (RAS) cells, whereas *CFTR* remarkable enriched in Ionocytes, AT2, RAS and secretory cells (also known as club cells) as well as in Mucous and Serous cells (**Figure 2A**). Co-expression of both mRNA was largely detected in AT2 cells (6.27%) with a few (greater than 1%) in RAS and ionocytes. Similarly, examination of the developing mouse lung at the embryonic and postnatal stages (23) revealed co-expression of *SLC26A9* and *CFTR* mRNA in the AT2, epithelial progenitor, and secretary (club) cells (**Figure 2B**).

**Figure 2.**
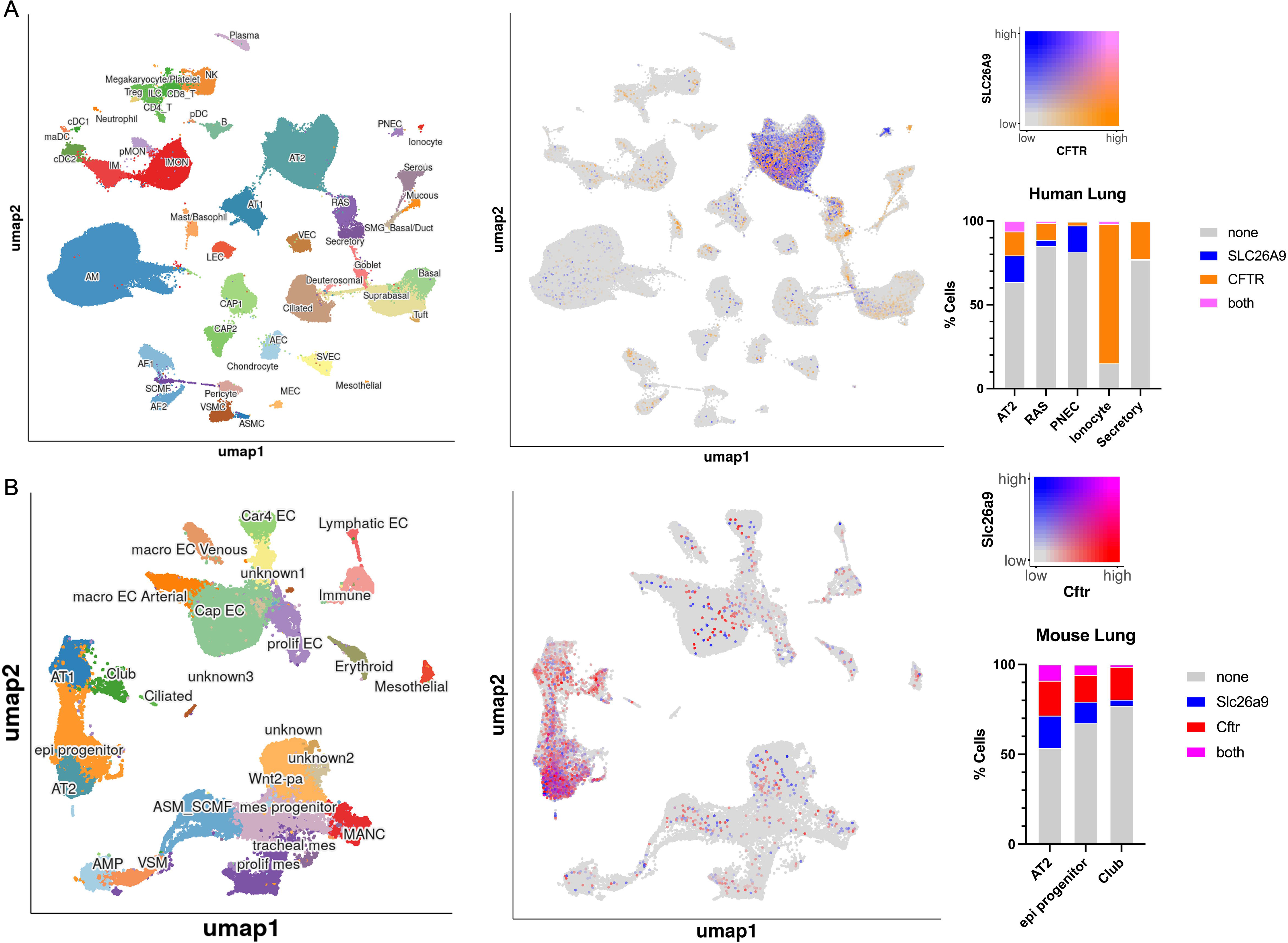
*SLC26A9* is co-expressed with *CFTR* in multiple cell types, mostly in Alveolar Type 2 (AT2), in both human **(A)** and mouse **(B)** lung tissues. The results shown here are in whole based upon data generated by the LungMAP Consortium [U01HL122642] and downloaded from (www.lungmap.net), on Sep 27, 2023. The LungMAP consortium and the LungMAP Data Coordinating Center (1U01HL122638) are funded by the National Heart, Lung, and Blood Institute (NHLBI).

### Structural Modeling for SLC26A9 variants

We summarized the RPCF-related *SLC26A9* exonic variants in **Table 2**, as well as their HGVS annotations with respect to hg19, MAFs in gnomAD, and mutation-effect predictions from CADD. These variants include NM_052934.4: c.229G>A, p.G77S (present in both RPCF1 and RPCF2); c.1885C>T, p.P629S (RPCF3); and NM_134325.2: c.2546G>A, p.R849Q (RPCF4). In addition, a synonymous variant NM_052934.4: c.966G>T, p.P322P was present in RPCF2.

**Table 2:** Summary of RPCF-related *SLC26A9* variants.

| Pt. no | Sex | Mutation Annotation<br>(Genomic, hg19) | Mutation Annotation<br>(Coding) | Mutation<br>Annotation<br>(Protein) | Mutation<br>Class | gnomAD MAF<br>[total count] | CADD-<br>PHRED |
| --- | --- | --- | --- | --- | --- | --- | --- |
| RPCF1 | M | chr1:205902109C>T | NM_052934.4 &<br>NM_134325.2<br>c.229G>A | p.G77S | Missense | 0.0004777<br>[282610] | 19.78 |
| RPCF2 | F |  |  |  |  |  |  |
| RPCF3 | M | chr1:205890864G>A | NM_052934.4 &<br>NM_134325.2 c.1885C>T | p.P629S | Missense | 0.0 | 17.29 |
| RPCF4 | F | chr1:205884138C>T | NM_134325.2 c.2546G>A | p.R849Q | Missense | 0.007357 [282862] | 2.819 |

We next sought to assess the location of these variants (G77S, P322P, P629S, and R849Q) within SLC26A9. SLC26A9 forms a homodimer (illustrated as molecule a and b) in the membrane to transport Cl- and Na+ (**Figure 3A**). Structural modeling shows that while P322P lies in an extracellular loop, G77 (colored in red) is located in the transmembrane 1 (TM1), which is part of the core region of the SulP transport domain and is close to the substrate sodium ion and the transport path indicated by the water molecules (**Figure 3B**). P629 is located at the loop in the cytoplasmic STAS domain, which has been shown potentially to interact with CFTR and to be involved with dimerization that facilitates SLC26A9 activity (24). The loop is too flexible to be identified in the cryo-EM structure. The two ends of the loop are colored in purple (**Figure 3C**). Both mutations G77S and P629S might affect the ion transportation capability of SLC26A9. The variant NM_134325.2: c.2546G>A, p.R849Q occurred in the isoform 2 extension, which was not captured by canonical isoform modeling.

**Figure 3:**
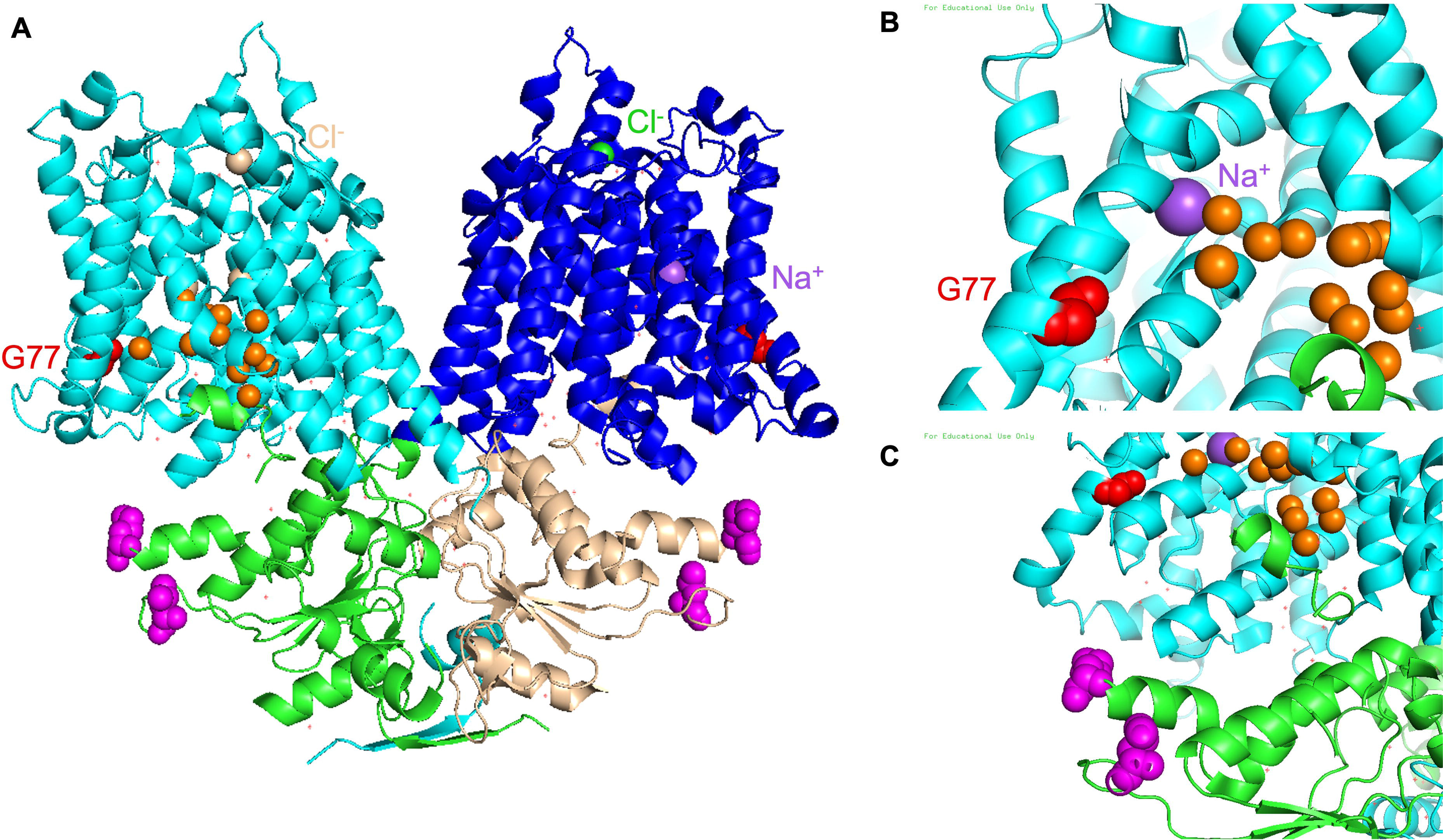
Molecular modeling of canonical isoform of SLC26A9. SLC26A9 forms a homodimer in the membrane to transport Cl- and Na+ (**A**). The transmembrane (TM) domain is colored in cyan and blue for molecule a and b, and the cytosolic STAS domain is colored in green and tint respectively. G77 (colored in red) is located at the transmembrane 1 (TM1), which is close to the substrate sodium ion and the transport path indicated by the water molecules (**B**). P629 is located at the loop in the cytoplasmic domain, STAS domain. The loop is too flexible to be identified in the cryo-EM structure. The two ends of the loop are colored in purple (**C**). Two of three exonic variants identified in our patients lie within the canonical isoform (NM_052934.4), which is modeled above, while p.R849 lies in the isoform 2 extension (NM_134325.2).

## Discussion

In patients with CF, defective CFTR disturbs the electrophysiological properties of the airway (1). In the lungs, disruption of CFTR leads to decreased chloride and bicarbonate secretion, and increased sodium and water absorption across the airway epithelial surfaces. This ion-transport defect results in a both acidic and dehydrated airway surface liquid (ASL) layer, inspissation of the mucus layer, defective mucociliary escalator function, and eventually a chronic vicious cycle of infection and inflammation leading to destruction of the lung tissue and ultimately respiratory failure (25, 26). The range of clinical signs and symptoms of lung disease in patients with CF is very broad. Various works have demonstrated that heritable factors other than *CFTR* may explain this variability (27). Here, we identified candidate modifiers using burden analysis and focused on modifiers that may exacerbate lung disease in patients with CF. To minimize bias, we performed the analysis on all coding genes rather than select genes and identified solute carrier family 26 member 9 (*SLC26A9*) as a modifier gene to be associated with RPCF.

SLC26A9 is an epithelial anion transporter that is preferentially expressed in the lung and gastrointestinal tract (28). In recent years, SLC26A9 has gained considerable attention, as genome-wide association studies (GWAS) demonstrated SLC26A9 as an important modifier of cystic fibrosis-related diabetes (CFRD), meconium ileus (MI), allergic airway disease, and response to new CF therapies (29-36). For example, the C allele of rs7512462 (chr1:205899595, GRCh37) in intron 5 of *SLC26A9*, which was first identified by GWAS of meconium ileus association in CF (29), is associated with greater expression of *SCL26A9* in the adult pancreas according to the Genotype Tissue Expression project (37) (GTEx; http://www.gtexportal.org/home/). The association of SLC26A9 SNPs with CF lung function is less clear. While one study questioned whether rs7512462 is linked with lung function (38), other studies have demonstrated a beneficial effect of the C allele on lung function or lung response to CFTR-modulators in individuals carrying CFTR variants (such as G551D) that result in better apical membrane localization for CFTR (31, 34). Individuals homozygous for F508del, comprising the majority of CF cases, showed a more modest association with rs7512462 (31), but the rescue of F508del-CFTR function by VX-809 and VX-770 in human nasal epithelial cells positively correlated with this SNP (39).

In this study, we report three *SLC26A9* missense variants (p.G77S, p.P629S, and p.R849Q) that were enriched in patients with RPCF carrying homozygous F508del mutations, but not found in patients with LTNP and controls. Cryo-EM structure analyses of murine and human SLC26A9 have revealed its unusual mode of oligomerization relying predominantly on the cytosolic STAS domain (18, 24). Structural modeling suggests that mutation (p.G77S) located in the SulP transport domain may disrupt its structure or process of ion transport across SLC26A9, whereas mutation (p.P629S) in the STAS domain may disrupt SLC26A9 homodimerization or its potential physical or functional interactions with CFTR, as previously reported of *SLC26A9* STAS-domain p.L683P and p.R575W mutation (40, 41). Mutations in the C terminal sequence as a gating modulator, however, may hamper its inhibitory effect on ion permeation. Structural modeling data supports that these missense variants in *SLC26A9* may lead to disrupted ion transport.

This study also demonstrated the feasibility of identifying, filtering, and collapsing variants as described above for a gene-based burden analysis to identify genetic modifiers in diseases such as CF. In this study, gene-based burden analysis provided greater power compared to SNP-based analysis, and Fisher’s exact test provided simple interpretability. Filtering out candidates that were also moderately associated with LTNP helped reduce incidental associations that may occur due to general association to CF or artifact. However, our approach may not adequately capture genes that have variants functioning in opposite directions, e.g. some variants associated with the rapid-progressing extreme of CF while other variants are associated with slowly progressing phenotype or no effect.

For decades, SLC26A9 is assumed to contribute to airway constitutive (basal) anion conductance regulated by CFTR (42, 43). It appears that SLC26A9 and CFTR behave differentially in polarized and non-polarized cells (44): SLC26A9 could stimulate CFTR activity by favoring the biogenesis and/or stabilization of CFTR in human bronchial cell lines (45), whereas CFTR interaction inhibits SLC26A9 anion transport activity when modeled in Xenopus oocytes (44, 46). Further studies showed that this constitutive anion secretion derived from SLC26A9 is absent in primary human bronchial epithelia (HBE) of CF donors (47). Similarly, HEK293 co-expressing SLC26A9 with the trafficking mutant F508del CFTR exhibited a significant reduction in constitutive current compared with cells co-expressing SLC26A9 and WT CFTR. SLC26A9 exhibited complex glycosylation when co-expressed with F508del CFTR, but its expression at the plasma membrane was decreased (47, 48).

However, recent results show that in both human airway epithelia and mouse trachea, basal and stimulated Cl^−^ secretion was due to CFTR despite clear expression of SLC26A9 in the apical membrane of ciliated cells (49). SLC26A9 may rather secrete HCO3^-^, thereby maintaining proper airway surface liquid (ASL) pH (49). It was speculated that depending on expression of CFTR, SLC26A9 may transport Cl^−^ or HCO3^−^□. Moreover, SLC26A9 does not secrete but probably supports reabsorption of fluid particularly in the alveolar space, which explains early death by neonatal distress in *Slc26a9*-knockout animals (50). Overall, these works indicate that *SLC26A9* and *CFTR* interplay and may have a complicated relationship depending on different tissues and cell types.

## Conclusion

In summary, we identify *SLC26A9* as one of the key CF modifier genes, by analyzing ES data from RPCF patients using an unbiased genome-wide genetic burden test. This finding supports SLC26A9 may act as an alternative ion channel in CF and compensate for CFTR defects in certain extent. *SLC26A9* deleterious mutations may abrogate this type of compensatory mechanisms and lead to more severe and rapid-progressing disease phenotype. However, there were limitations to our study highlighting the need to study *SLC26A9* further in the context of extreme lung disease. Gaining a deeper insight into *SLC26A9* as a potential modifier for CF may facilitate personalized medicine.

## Acknowledgements

The authors would like to thank the patient and their parents for their commitment to this research and Casie Genetti for enrollment of those families to the study. The Genotype-Tissue Expression (GTEx) Project was supported by the Common Fund of the Office of the Director of the National Institutes of Health, and by NCI, NHGRI, NHLBI, NIDA, NIMH, and NINDS. The data used for the analyses described in this manuscript were obtained from the GTEx Portal on 10/10/23.

## Author contributions

PBA designed the experiments and project administration. All the authors participated in performing the experiments, data analyses, manuscript drafting and/or figure generation. All the authors read and approved the final manuscript.

## Data Availability Statement

Research data are stored in an institutional repository and will be shared upon request to the corresponding author.

